# Alterations in Aortic Elasticity Indices among Type 2 Diabetes Patients in a Low and Middle Income Country Using M-Mode Echocardiography: A Cross-Sectional Comparative Study

**DOI:** 10.1101/2024.06.06.24308579

**Authors:** Hai Nguyen Ngoc Dang, Thang Viet Luong, Quan Nguyen Khoi, Uyen Ngoc Phuong Nguyen, Nguyen Nguyen Khoi Pham, Hieu Thi Nguyen Tran, Hung Khanh Tran, Mai Thi Thu Cao, Binh Anh Ho, Thang Chi Doan, Hung Minh Nguyen, Tien Anh Hoang, Minh Van Huynh

**Author notes:** Hai Nguyen Ngoc Dang and Thang Viet Luong are co-first authors and contributed equally to this work. Author contributions: Conceptualization: Hai Nguyen Ngoc Dang, Thang Viet Luong, Quan Nguyen Khoi, Nguyen Nguyen Khoi Pham. Data curation: Hai Nguyen Ngoc Dang, Thang Viet Luong. Formal analysis: Hai Nguyen Ngoc Dang, Thang Viet Luong. Investigation: Hai Nguyen Ngoc Dang, Thang Viet Luong. Methodology: Hai Nguyen Ngoc Dang, Thang Viet Luong. Supervision: Hai Nguyen Ngoc Dang, Thang Viet Luong. Writing – original draft: Hai Nguyen Ngoc Dang, Thang Viet Luong, Quan Nguyen Khoi, Uyen Ngoc Phuong Nguyen, Nguyen Nguyen Khoi Pham, Hieu Thi Nguyen Tran, Hung Khanh Tran, Mai Thi Thu Cao, Binh Anh Ho, Thang Chi Doan, Hung Nguyen Minh, Tien Anh Hoang, Minh Van Huynh. Writing – review & editing: Hai Nguyen Ngoc Dang, Thang Viet Luong, Quan Nguyen Khoi, Uyen Ngoc Phuong Nguyen, Nguyen Nguyen Khoi Pham, Hieu Thi Nguyen Tran, Hung Khanh Tran, Mai Thi Thu Cao, Binh Anh Ho, Thang Doan Chi, Hung Nguyen Minh, Tien Anh Hoang, Minh Van Huynh. All authors have read and agreed to the published version of the manuscript.

## Abstract

**Background:** Diabetes is one of the leading causes of noncommunicable diseases worldwide. It is known to induce cardiovascular remodeling, which can result in a variety of complications, including a considerable increase in aortic stiffness. While studies in Western populations have explored these effects, data on Asians, particularly Vietnamese, are limited. This study aimed to assess aortic elasticity in type 2 diabetes mellitus (T2DM) patients compared to healthy individuals.

**Methods:** Aortic elasticity indices were measured using M-mode echocardiography. A comparison between the healthy group and the T2DM group revealed substantial differences in aortic elasticity indices.

**Results:** The aortic stiffness index (ASI) was significantly greater in the T2DM group than in the control group, with median values of 6.10 (3.64-12.47) and 3.79 (2.40-8.50), respectively (p = 0.003). Aortic strain (AS) was substantially lower in the T2DM group than in the control group, with median values of 8.21 (4.24 - 13.07) and 10.66 (6.01 - 18.23), respectively (p = 0.039). Furthermore, the median aortic compliance (AC) and aortic distensibility (AD) in individuals with T2DM were 4.07 (2.28-7.44) and 3.08 (1.57-5.26), respectively, lower than those in the control group, with median values of 6.40 (3.08-10.75) and 5.33 (2.80-9.79), respectively. A longer diabetes duration was linked to a greater ASI (r = 0.43, p < 0.05), while the AS decreased (r = -0.37, p < 0.05).

**Conclusions:** Substantial variations in aorta elasticity indices were found using M-mode echocardiography. More research is needed to investigate the consequences of these discrepancies and their significance for clinical purposes.

**Graphical Abstract:** 

## Introduction

Type 2 diabetes mellitus (T2DM) is currently an emerging noncommunicable illness worldwide. According to the International Diabetes Federation (IDF), the number of individuals with T2DM is expected to increase globally, with 10.5% of people aged 20 to 75 years being diagnosed with diabetes by 2021. By 2045, this figure will increase by 46%, with an estimated 783 million people living with diabetes. Diabetes caused 2.3 million fatalities in the Western Pacific region in 2021, the highest among all IDF regions. Among these, Vietnam has almost 57 thousand diabetes-related fatalities in adults aged 20 to 79 years [1].

Many studies have shown that T2DM patients experience changes in both cardiac morphology and function over time after being diagnosed with the condition [2,3]. In addition to damaging the heart, vascular remodeling has recently been shown to occur in patients with diabetes patients. Numerous studies on diabetes mellitus have revealed a strong link between vascular remodeling and disease, which is affected by a variety of variables [4–6]. The most important factors are insulin resistance and dyslipidemia [7]. Furthermore, because diabetes mellitus involves the risk of persistent hyperglycemia, glucose creates covalent connections with proteins, nucleic acids, and lipids by nonenzymatic glycosylation, also known as glycation. This irreversible process results in advanced glycation end products (AGEs). These chemicals accumulate in the vascular wall and, with time, impair the flexibility of blood vessels [8]. As diabetes progresses, its complications may worsen. The duration of the condition (since first diagnosis) can be seen as the natural progression of vascular complications that are also associated with an increased risk of death from coronary artery disease or stroke due to structural abnormalities in the cardiovascular system. There are several invasive and noninvasive procedures for measuring vascular elasticity. These include pulse pressure, invasive blood pressure, and monitoring variations in blood vessel size. Imaging techniques such as magnetic resonance imaging, computed tomography, and ultrasound provide noninvasive but effective methods for measuring arterial elasticity. Among these approaches, ultrasound stands out for its low cost, ubiquitous availability, and broad applicability, making it the recommended tool for measuring arterial elasticity.

Aortic elasticity, a measure of arterial stiffness, has been demonstrated to be a strong independent predictor of atherosclerosis and other cardiovascular events. This shows that aortic flexibility could be a useful technique for detecting vascular issues in their early stages [9]. However, most previous studies have concentrated on Western or Caucasian populations. There is a considerable knowledge gap about how aortic elasticity may change in people with T2DM in populations of low- and middle-income countries in general and specifically in the population of Vietnam. This study was conducted to investigate the indications of aortic elasticity in T2DM patients and compare the aorta elasticity indices of T2DM patients and healthy control participants to determine patterns of change. By analyzing these parameters, this study aimed to gain valuable insights into how T2DM affects cardiovascular health, especially aortic elasticity, and contributes to an effort to detect and reduce disease burdens globally early.

## Methods

### Study population

This is a quantitative, comparative cross-sectional study with a comparison group. We integrated components from the STROBE declaration to enhance the quality of reporting observational studies [10]. This study included 109 subjects who underwent echocardiography at the Cardiac Ultrasound Unit of the Hue University of Medicine and Pharmacy from 15/04/2022 to 01/06/2023. The study received approval from The Institutional Ethics Committee of Hue University of Medicine and Pharmacy (Approval number: H2022/025) and was conducted under the principles outlined in the Declaration of Helsinki 2013. Prior to participation, written informed consent was obtained from each patient or their legal representative.

Among the 109 participants, 47 were healthy subjects who underwent regular health check-ups, had no history of diabetes or cardiac pathology, and were currently free from cardiac or diabetic conditions. Another 62 individuals were diagnosed with T2DM and had no known comorbidities according to the diagnostic criteria stipulated by the American Diabetes Association in 2022 [11].

Patients who refused to participate in the study, as well as those with known comorbidities such as hypertension, were excluded. Patients with any other cardiovascular disease (severe aortic stenosis, moderate to severe aortic regurgitation, severe mitral valve stenosis, congenital heart disease, cardiomyopathy, myocardial ischemia, reduced ejection fraction, arrhythmia, or chronic atrial fibrillation) were also excluded. We also excluded patients who had pacemakers inserted, pregnant or breastfeeding women, people who suffered from acute life-threatening medical illnesses, and those who had hazy or inadequate ultrasound images.

### Data collection

The data were collected using an ultrasound machine, paper medical records, and data collection forms to extract information from the study subjects. This process involved the following steps:

### Clinical examination and blood tests

After consenting to participate in this study, the patient’s medical history was obtained, with an emphasis on characteristics such as the time of T2DM diagnosis and blood sugar management at each visit.

Body mass index (BMI) was computed using the following formula: BMI = weight (kg)/[height (m) × height (m)]. Body surface area (BSA) was calculated using the formula of Du Bois [12]: BSA = 0.007184 × (weight)^0.425^ × (height)^0.725^. Blood pressure was measured following the guidelines of the American Heart Association in 2019 [13,14].

Blood samples for biochemical analysis were collected from fasting venous blood after waking. Biochemical tests were performed on a Roche Cobas E601 automated clinical chemistry analyzer at the Central Testing Unit of Hue University of Medicine and Pharmacy Hospital.

### Transthoracic echocardiography

Our study utilized a specialized Affiniti 70 echocardiography machine from Philips, the Netherlands, with the ability to perform time-motion mode (M-mode), two-dimensional mode, continuous-wave Doppler, color Doppler, and tissue Doppler imaging. The ultrasound machine simultaneously recorded the electrocardiography data along with the echocardiographic images. Transthoracic echocardiography was performed following the American Society of Echocardiography guidelines for adults [15].

### Arterial stiffness indices

M-mode echocardiography recorded the diameter of the ascending aorta 3 cm above the aorta valve in a parasternal long-axis image.

The end-diastolic diameter of the ascending aorta (AoD) was measured at the peak of the R wave on the ECG, and the end-systolic diameter of the ascending aorta (AoS) was measured at the maximum anterior motion of the aorta (see **Fig 1**). The diameter of the ascending aorta was measured in centimeters (cm). Arterial stiffness was assessed using five indices: aortic strain (AS) % = 100 x (AoS – AoD)/AoD, aortic stiffness index (ASI) = ln (SBP/DBP)/[(AoS – AoD)/AoD], aortic compliance (AC) mm/mmHg = [10 x (AoS – AoD)]/(SBP – DBP), aortic distensibility (AD) ‰mmHg^−1^ = [1000 x 2 x (AoS – AoD)]/[AoD x (SBP – DBP)], and aortic Peterson’s elastic modulus (AEM) mmHg = [(SBP – DBP) x AoD]/(AoS – AoD). Note: AoS refers to the aortic end-systolic diameter; AoD denotes the aortic end-diastolic diameter; SBP represents systolic blood pressure; DBP stands for diastolic blood pressure [16–18].

**Fig 1.**
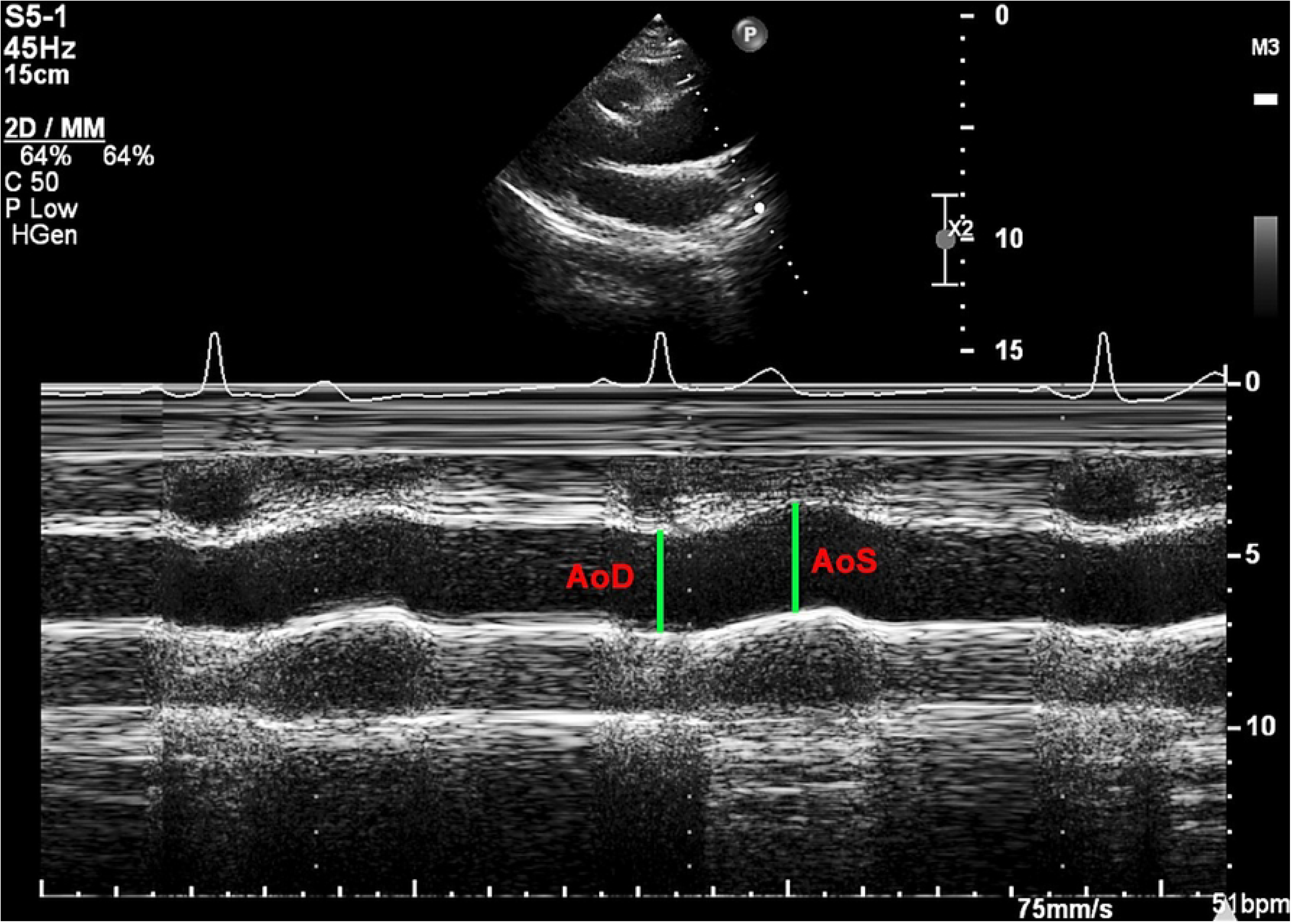
The aorta was visualized in M-mode. AoD: end-diastolic diameter of the ascending aorta; AoS: end-systolic diameter of the ascending aorta.

### Statistical analysis

The statistical analysis was carried out using SPSS software version 26 (IBM, Armonk, NY, USA). The Kolmogorov‒Smirnov test was used to determine the normality of the data. Normally distributed continuous variables are reported as the means ± standard deviations. Variables that were not normally distributed are represented as medians with interquartile ranges (IQRs = 25^th^ – 75^th^ percentiles). Categorical variables are represented as frequencies and percentages. Missing data were omitted from the analyses. Individuals with and without T2DM were compared based on their baseline characteristics. Continuous variables were analyzed using independent t tests, and the results are reported as the means ± standard deviations. The Mann‒Whitney nonparametric test was applied for the analysis of variables that were not normally distributed. Categorical variables are presented as n (%) and were compared using the chi-squared test.

To compare arterial elastic parameters between the T2DM and non-T2DM groups, the Mann‒Whitney test was used for nonnormally distributed continuous variables. Spearman’s correlation coefficient (r) and its related P value elucidated the correlation between continuous variables. All probability values were two-sided, and P values less than 0.05 were considered to indicate statistical significance. We followed the SAMPL guidelines in our statistical analysis to prevent avoidable errors or omissions in reporting statistical data [19].

## Results

### Clinical and echocardiographic features

This study included 109 subjects, including 62 T2DM patients and 47 healthy individuals who met the diagnostic criteria and composed the comparison group. Our research revealed the following results.

There were no significant differences in age, sex, height, weight, BMI, or BSA between the groups (p > 0.05). The differences in the left atrial diameter (LAd), left ventricular mass index (LVMI), and ejection fraction (EF) between the T2DM group and control group were not statistically significant (p > 0.05). Detailed information regarding these indices is presented in **Table 1**.

**Table 1.** Anthropometric and echocardiographic characteristics of participants by group.

| Characteristics | T2DM group<br>(n = 62) | Comparison group<br>(n = 47) | P value |
| --- | --- | --- | --- |
| Age (years) | 61.63 ± 10.46 | 60.40 ± 7.08 | 0.491 |
| **Female (%)** | 40 (64.5%) | 26 (55.3%) | 0.561 |
| **BMI (kg/m<sup>2</sup>)** | 22.26 ± 3.06 | 22.49 ± 2.35 | 0.657 |
| **BSA (m<sup>2</sup>)** | 1.53 ± 0.15 | 1.56 ± 0.13 | 0.300 |
| **HbA1C (%)** | 10.92 ± 3.02 | 5.03 ± 0.38 | <0.001 |
| **Duration of diabetes (years)** | 6.00 [4.00 – 14.25] | NA |  |
| **SBP (mmHg)** | 127.63 ± 8.85 | 115.1 ± 12.19 | <0.001 |
| **DBP (mmHg)** | 72.63 ± 7.02 | 72.32 ± 7.59 | 0.824 |
| **LAd (cm)** | 3.15 ± 0.42 | 3.19 ± 0.45 | 0.645 |
| **LVMI (g/m<sup>2</sup>)** | 107.31 ± 30.39 | 103.58 ± 23.08 | 0.488 |
| **LVEF (%)** | 67.92 ± 7.53 | 70.23 ± 5.27 | 0.075 |
| **Total cholesterol**<br>**(mmol/L)** | 4.97 [3.63 – 6.49] | 4.98 [4.15 – 5.00] | 0.503 |
| **Triglyceride**<br>**(mmol/L)** | 1.85 [1.06 – 2.95] | 1.20 [1.05 – 1.25] | 0.001 |
| **HDL-C (mmol/L)** | 0.96 [0.81 – 1.22] | 1.25 [1.22 – 1.30] | < 0.001 |
| **LDL-C (mmol/L)** | 3.13 [1.96 - 3.67] | 3.45 [2.73 – 3.55] | 0.264 |

The values are presented as the means ± standard deviations, medians [Q25 - Q75] or numbers (%). T2DM: type 2 diabetes mellitus; BSA: body surface area; BMI: body mass index; SBP: systolic blood pressure; DBP: diastolic blood pressure; LAd: left atrial diameter: LVMI: left ventricular mass index; LVEF: left ventricular ejection fraction; HbA1c: glycated hemoglobin; HDL-C: high-density lipoprotein cholesterol; LDL-C: low-density lipoprotein cholesterol.

### Aortic elasticity indices in the T2DM and control groups

The T2DM group had significantly lower AC, AD, and AS than did the control group. The T2DM patients also had significantly greater ASI and AEM than did the controls (p < 0.05). Additionally, the control group had a greater AS of 10.66 (6.01 - 18.23) than did the T2DM group at 8.21 (4.24 - 13.07). Detailed information is provided in **Table 2** and **Fig 2**.

**Fig 2.**
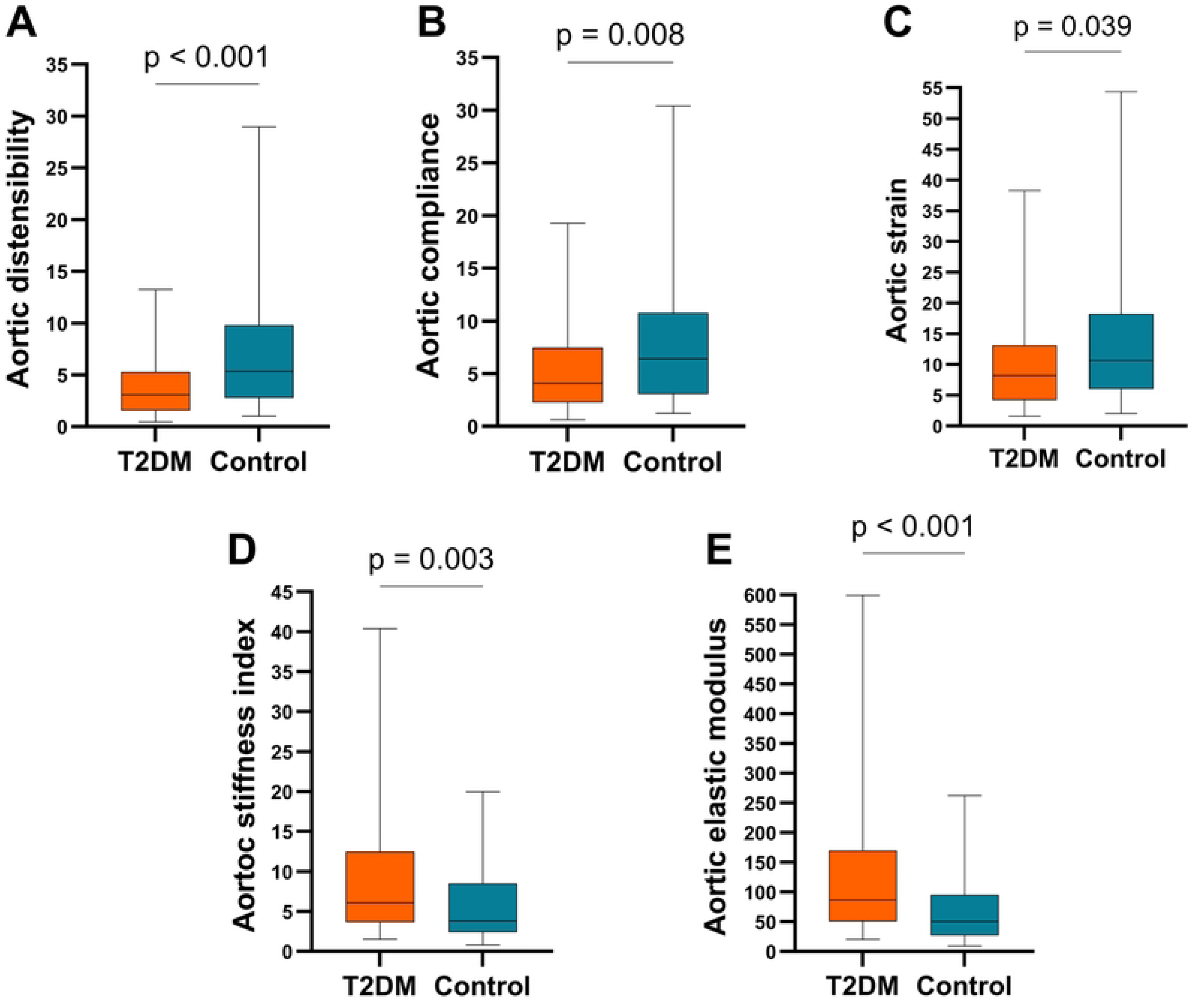
Comparison of aortic elasticity indices. (A) Comparison of aortic distensibility between the diabetes group and control group. (B) Comparison of aortic compliance between the diabetes group and the control group. (C) Comparison of strains between the diabetes group and control group. (D) Comparison of the aortic stiffness indices between the diabetes group and the control group. (E) Comparison of Peterson’s elastic modulus between the diabetes group and control group. T2DM: diabetes mellitus.

**Table 2.**
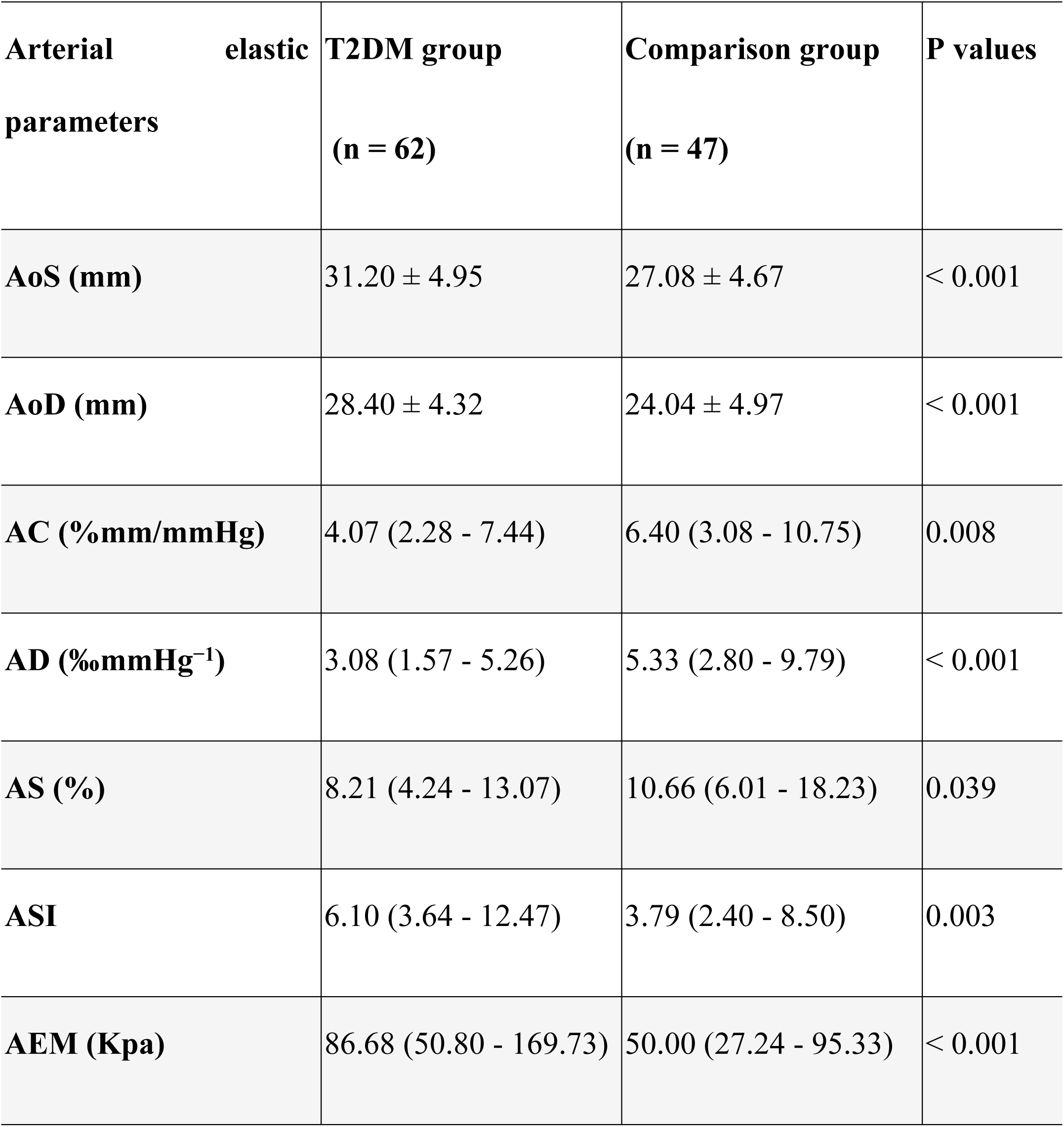
Computed aortic elasticity parameters of participants by group.

The values are presented as the means ± standard deviations or medians [Q25 - Q75]. T2DM: Type 2 diabetes mellitus; AoS: aortic end-systolic diameter; AoD: aortic end-diastolic diameter; AC: aortic compliance; AD: aortic distensibility; AS: aortic strain; ASI: aortic stiffness index; AEM: Aortic Peterson’s elastic modulus

### Correlations between indices of aortic elasticity and relevant variables

An analysis was performed to explore the relationships between anthropometric and echocardiographic features and computed aortic indices. **Fig 3** visually illustrates the results in a heatmap. The results revealed that in the T2DM group, the duration of diabetes was associated with arterial elasticity indicators. A longer duration of diabetes was positively associated with greater ASI and AEM (Spearman’s correlation coefficients of 0.43 and 0.43, respectively). In contrast, AC, AD, and AS tended to decrease as diabetes duration increased (Spearman’s correlation coefficients of -0.39, -0.43, and -0.37, respectively).

**Fig 3.**
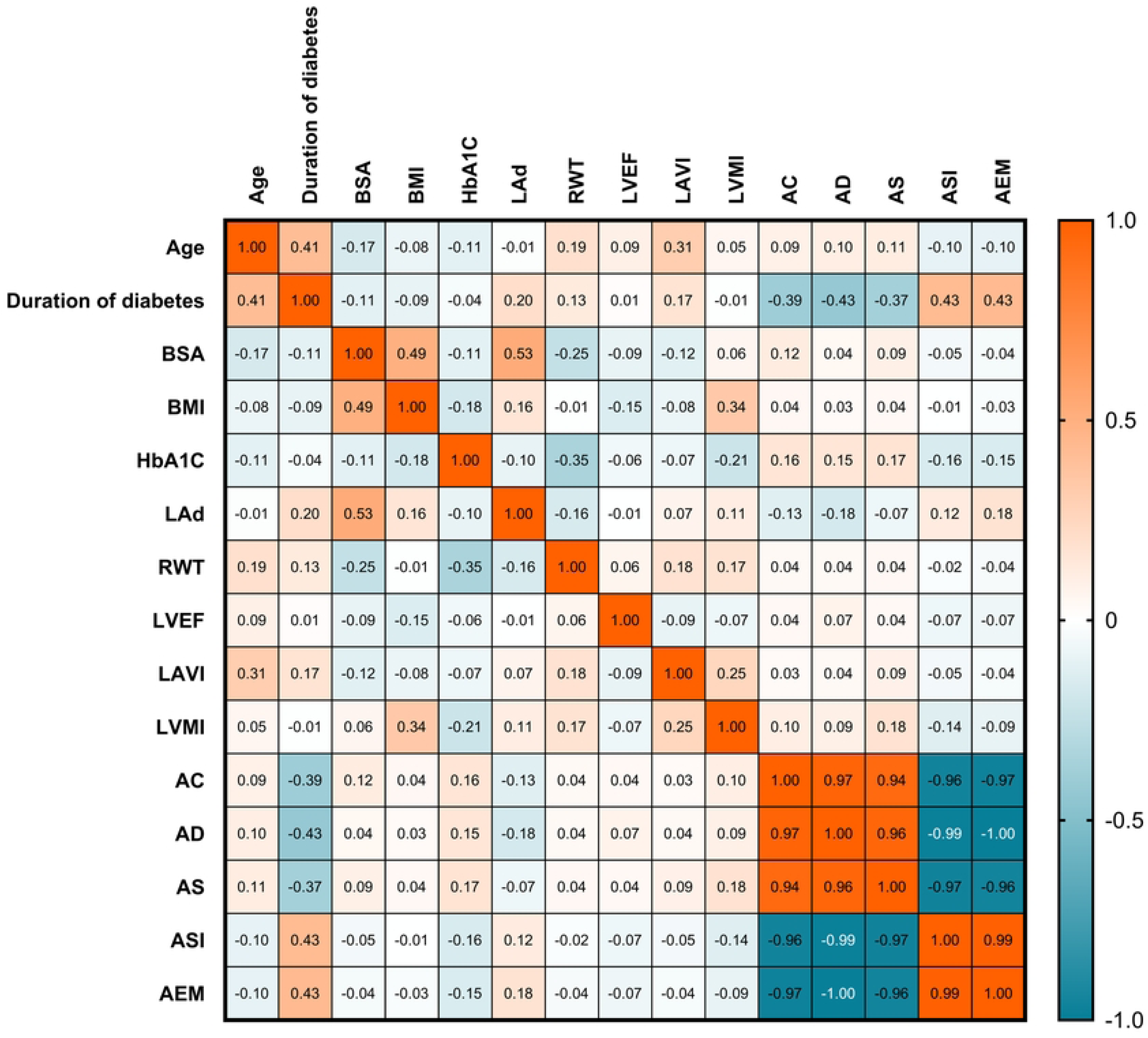
Correlations between anthropometric and echocardiographic features and computed aortic indices. BSA: body surface area; BMI: body mass index; HbA1c: glycated hemoglobin; LAd: left atrial diameter; RWT: relative wall thickness; LVEF: left ventricular ejection fraction; LVMI: left ventricular mass index; AC: aortic compliance; AD: aortic distensibility; AS: aortic strain; ASI: aortic stiffness index; AEM: Aortic Peterson’s elastic modulus.

## Discussion

There is growing interest in studying the association between aortic flexibility and cardiovascular disease. In contrast, the stage of myocardial performance in diabetes and prediabetes patients remains overlooked [20]. Among the wide range of choices for echocardiography, M-mode echocardiography is still effective at capturing the motions and functions of the heart and surrounding structures. Despite the better temporal resolution of M-mode echocardiography, it is still being disregarded in international guidelines [21]. Substantial efforts have been made to investigate the relationships between insulin resistance (and T2DM) and aortic elastic characteristics. This study was performed in Vietnam with a group of T2DM patients from low- and middle-income countries to assess the feasibility and cost-effectiveness of M-mode echocardiography in daily practice.

### Comparison of aortic elasticity indices between patients with T2DM and healthy controls

It is generally known that vascular elasticity often begins to decline in middle age and worsens over time [17]. Throughout the aging process, the elasticity of the endothelium layer gradually decreases, which has a significant impact on the structure of the arteries due to reduced elastic fiber activity, particularly in major arteries such as the aorta [22]. The deterioration of arterial elasticity is caused by a variety of mechanisms, including the impairment of vasodilation mechanisms via nitric oxide (NO) and the absence of ETB receptors, which increase the plasma endothelin-1 concentration because these receptors contribute to endothelin-1 destruction [23,24].

In this study, we measured the elasticity of the aorta using five indices: AC, AD, ASI, AS, and AEM. The T2DM group had considerably lower AC, AD, and AS indices than did the comparison group but had higher ASI and AEM. This study successfully reproduced the findings previously reported by Malgorzata Dec-Gilowska et al [17]. Furthermore, our findings are consistent with those of Liu et al., who conducted a study on individuals with diabetes patients and concluded that these patients are more likely to experience arterial stiffness than healthy individuals are [25]. According to Zapolski et al., patients with diabetes have greater ASI readings than healthy people, independent of whether their disease is accompanied by hypertension [26]. Another study by Aslan also confirmed that patients with prediabetes have a greater ASI than healthy people according to echocardiography [20].

An increased ASI is an independent risk factor for cardiovascular disease and mortality [27]. Our findings are consistent with those of Dec-Gilowska et al., who reported that diabetes patients tend to have a significantly decreased AC. In parallel, our investigation revealed a rise in ASI akin to the observations made by Dec-Gilowska et al [28]. We found that both the ASI and the AEM nearly doubled in patients with T2DM compared to those in the control group. It is well known that the ASI is strongly correlated with the mean arterial pressure, and increased ASI may account for the development of hypertension. However, this study is unable to shed light on the question of whether either ASI or hypertension precedes the other in this progression among T2DM patients.

Despite major breakthroughs in T2DM treatment, the risk of cardiovascular morbidity and mortality continues to increase [29]. Recent research has revealed a complex link between cardiovascular mortality risk and T2DM, which is thought to be caused in part by increased arterial stiffness [30]. The literature has shown that people with T2DM have a much greater incidence of increased arterial stiffness than individuals without diabetes [31]. This can be due to insulin resistance, hyperglycemia, and persistent hyperinsulinemia. These drugs activate the renin-angiotensin-aldosterone system and enhance the expression of type 1 angiotensin receptors in vascular tissue, inducing arterial wall hypertrophy and fibrosis [32–34]. Insulin resistance inhibits PI3-kinase-dependent enzyme signaling, which is involved in insulin metabolism, cell growth, proliferation, differentiation, motility, survival, and intracellular transport, resulting in hyperinsulinemia. Furthermore, decreased glucose tolerance increases glycation and changes the mechanical characteristics of the artery wall’s interstitial tissue via a variety of complex mechanisms [35,36]. Increased arterial stiffness causes irreversible pathogenic loads, particularly in patients with diabetes. A constant increase in arterial stiffness results in increased transfer of pulse pressure into the microcirculation, which is directly associated with a greater risk of disorders such as hypertension, chronic renal disease, coronary artery disease, heart failure, and stroke [37–40]. Our study revealed substantial differences in AC, AD, AS, ASI, and AEM between individuals with T2DM and individuals without diabetes, which suggests further screenings and interventions for these patients are needed for better outcomes.

### Correlations between aortic elasticity indices and diabetic conditions

Our study did not identify any correlation between the HbA1c level and aortic elasticity indices (**Fig 3**).

The association between HbA1c, an indicator of long-term blood glucose regulation, and aortic stiffness seems to be constrained in certain studies [41–43]. This constraint can be explained by the fact that HbA1c measurements reflect only a snapshot of glucose levels attached to hemoglobin over the preceding three months rather than the comprehensive control of blood sugar over an extended period. Consequently, it may not accurately correlate with the development of arterial stiffness, which occurs gradually over time [44].

Emerging evidence suggests that blood glucose variability could exert a more significant impact on aortic stiffness. Individuals with diabetes, particularly those with poorly managed blood glucose levels and significant variability, face an elevated risk of aortic stiffness and related cardiovascular complications [45,46]. Therefore, emphasizing the control of blood glucose fluctuations through comprehensive diabetes management strategies is pivotal for mitigating the risk of aortic stiffness and enhancing cardiovascular health outcomes among individuals with diabetes.

Nevertheless, we did find significant correlations between the duration of diabetes and arterial elasticity indices as shown in the heatmap (**Fig 3**). A longer duration of diabetes is linked to greater stiffness, whereas elasticity decreases accordingly.

In diabetes, the duration of T2DM is intricately linked to the progression of disease complications [47]. In a study led by Agnoletti et al., a significant correlation emerged between the duration of diabetes and the worsening of arterial stiffness. Their meticulous findings revealed that the duration of diabetes correlates with microvascular complications, regardless of renal function, and with macrovascular complications due to the presence of increased aortic stiffness [48]. By recognizing the pivotal role of diabetes duration in the progression of arterial stiffness, early diagnosis, and proactive treatment become paramount in optimizing patient care and outcomes.

### Limitations of the study

The single-center design and limited sample size constrain our study. The small sample size is particularly restrictive, potentially introducing bias and diminishing statistical power. Therefore, there is a need for multicenter research with a larger sample size to obtain more vital clinical evidence. Additionally, our study is only a cross-sectional investigation that can demonstrate associations but not causal relationships. Hence, longitudinal studies are required to address this issue. Furthermore, arterial stiffness parameters were measured in the central aorta, whereas blood pressure was measured in peripheral arteries. However, due to ethical considerations, our study solely relied on peripheral blood pressure measurements instead of invasive arterial pressure measurements. Additionally, given the circumstances at our research site, where alterations in medication regimens are common, our study did not assess the impact of different drug treatments on arterial stiffness. Thus, more significant and longer-term studies are needed to evaluate this aspect.

## Conclusions

A notable difference in aortic elasticity indices was detected between the T2DM patient group and the control group via M-mode echocardiography.

## Data Availability

Data cannot be shared publicly due to certain local ethical constraints. Researchers who meet the criteria for access to confidential data may contact the author, Hai Nguyen Ngoc Dang (via email at)

